# High burden of chronic kidney disease of unknown cause among patients receiving renal replacement therapy in Northeast Nigeria: A cross-sectional survey of haemodialysis units

**DOI:** 10.1101/2024.06.24.24309383

**Authors:** Baba Waru Goni, Hamidu Suleiman Kwairanga, Aliyu Abdu, Ibrahim Ummate, Alhaji Abdu, Ahmed Ibrahim Ba’aba, Mohammad Maina Sulaiman, Loskurima Umar, M.L Gana, Aliyu Abdulkadir, Sabiu Musa, Shatuwa Adamu, Idris A. Usman, Hauwa Alhaji Sabo, Idris Musa Abubakar, Hamza Bukar Adam, Ismail Alhaji Umar, Ayabaryu Papka, Modu Mustapha, Yauba Mohammed Saad, Amin Oomatia, Mahmoud Bukar Maina, Neil Pearce, Ben Caplin

## Abstract

**Introduction:** Chronic kidney disease (CKD) is emerging as a significant public health concern in northeastern Nigeria, particularly in states such as Yobe and Borno. Despite its increasing impact, there is a lack of data characterizing this public health issue. This study aims to explore the prevalence, spatial distribution, and risk factors for CKD among patients receiving haemodialysis (HD) in the region.

**Methodology:** A cross-sectional survey of HD centres in Yobe, Borno, and Jigawa States of Nigeria was conducted. Questionnaire responses were obtained on demographic, social, and clinical data. Spatial analyses were conducted to determine the geographic distribution of the cases.

**Results:** We identified 376 patients receiving HD services across 4 centres. Of these, 207 (55.1%) were male and the mean age was 46.56 ± 16.4. Most patients reside in urban areas (67.6%). The main pre-dialysis occupations included civil service (100 [26.6%]), agriculture (65 [17.3%]), and trading (58 [15.4%]). ‘Hypertension’ (195 [51.9%]) was the most common self-reported primary renal disease, followed by unknown causes (70 [18.6%]) and Diabetic Kidney Disease (30 [8%]). Regional analysis demonstrated a particularly high burden of disease in Bade and Jakusko Local Government Areas.

**Discussion and Conclusion:** Spatial analysis suggests the existence of a CKD hotspot geographically associated with communities along the River Yobe, raising the possibility of an important environmental cause of disease. This study also highlights the lack of access to adequate diagnosis and geographical clustering of CKD burden in this region. These findings further reinforce the need for population-representative studies to characterize the burden of CKD alongside strategic healthcare interventions and collaboration among stakeholders aimed at improving access to care.

## INTRODUCTION

CKD is a significant global health issue with an estimated prevalence of 13.4%. ^1,2^ Its emergence as a major contributor to morbidity and mortality worldwide highlights the significance for comprehensive research and intervention across populations. ^1,3^ The global rise in CKD is attributed to factors such as diabetes mellitus, hypertension, obesity, and an aging population^4^. However, regional variations exist, with unique determinants like infections and exposure to environmental toxins contributing to CKD’s prevalence in specific areas^5^.

CKD of unknown cause (CKDu) has been reported in rural communities of developing countries, including Sri Lanka, India, Central American nations, and North Africa ^1^. Unlike typical CKD, CKDu is not linked to common risk factors such as hypertension or diabetes. In Nigeria, particularly in the North Eastern region along the Kumadugu River valley, anecdotal reports and small-scale studies have indicated a high prevalence of CKD. ^1,3^ The Bade community in Northern Yobe State has been identified as a CKD hotspot, with a significant number of CKD cases without a clear underlying cause.^6^

To bridge this knowledge gap, we conducted a cross-sectional survey of HD centers in the region. This approach aligns with the ‘passive detection’ strategies recommended by the International Society of Nephrology’s International Consortium of Collaborators on CKDu.^7^ The survey aims to delineate the geographic distribution, underlying diagnoses, and risk factors for disease among the HD population in Yobe and surrounding states.

## METHODS

### Data Collection

Ethical approval for the study was obtained from the study centres, as well as the Yobe State Ministry of Health in Nigeria. Data collection for this study was carried out from January to June 2023, coinciding with patients’ scheduled HD sessions at designated centers to ensure minimal disruption to their routine care. The study encompassed all HD facilities in the north-eastern states of Borno and Yobe, specifically Yobe State University Teaching Hospital (YSUTH) in Damaturu, Federal Medical Center in Nguru (FMC_Nguru), University of Maiduguri Teaching Hospital (UMTH), and State Specialist Hospital in Maiduguri (SSH_Maiduguri). Additionally, State Specialist Hospital Hadejia (SSH_Hadejia) in Jigawa State was included to provide a broader perspective on the CKD prevalence in the region. Despite Jigawa’s location in northwestern Nigeria, Hadejia is the largest town in the north-eastern part of Jigawa state potentially providing treatment for patients living in Yobe state.

Patients were individually approached for participation, and informed consent was secured from each before inclusion in the study. Trained data collectors then administered a comprehensive, researcher-adapted questionnaire to those who consented. This questionnaire was designed to capture a range of information, including demographic details (age, gender), clinical data (duration and frequency of dialysis treatments), and potential factors contributing to the etiology of CKD. Additional relevant variables, such as occupation and residential history, were also gathered to support a multifaceted analysis of CKD within the population.

## Data Analysis

Data analysis was performed using Python 3.10. Descriptive statistics in the form of mean (SD), median (IQR) and frequencies were used to characterize the study population. Geographical data was presented using geopandas and geoplots for mapping HD prevalence across the study region. Data visualizations, including bar charts and heatmaps, were created using matplotlib and seaborn.

## RESULTS

### Description of study population by centre

The study enrolled 376 participants (Table 2), with a higher proportion of males (55.1%) than females (44.9%). Variations in sex distribution were observed among centers, with YSUTH having the highest male representation (61.6%) and FMC_Nguru reporting the highest female representation (54.5%). The mean age was 46.56 ± 16.4 years, varying across centers. SSH Maiduguri and UMTH had the highest urban representation (91.9% and 75.6% respectively). Civil servants comprised 26.6% of participants, with SSH_Maiduguri reporting the highest proportion (38.4%). Traders accounted for 15.4%, again with SSH_Maiduguri reporting the highest (27.3%). The agriculture sector represented 17.3%. Transport and fishing occupations were less represented, and no participants were from the mining sector.

**Table 1:** Inclusion And Exclusion Criteria.

| Inclusion Criteria |  |
| --- | --- |
| 1. | Individuals receiving HD treatment at the selected HD centers between January 2023 and June 2023 |
| 2 | Patients who were diagnosed with established CKD. |
| 3 | Patients who provided informed consent to participate in the study. |
| Exclusion Criteria |  |
| 1 | Patients undergoing HD treatment outside the designated study centers even if they were living in the state under study. |
| 2 | Individuals with acute kidney injury (AKI), which is defined as an abrupt decrease in kidney function that includes both injury (structural damage) and impairment (loss of function). |
| 3 | CKD patients who are not on HD |
| 4 | Those who did not provide informed consent. |
| 5 | CKD patients receiving HD services at more than one centre. |

**Table 2:** Description of study population by centre.

| Table 2: Description of study population by centre |  |  |  |  |  |  |  |
| --- | --- | --- | --- | --- | --- | --- | --- |
| Variables |  | All centres | ysuth | umth | ssh_maiduguri | fmc_nguru | ssh_hadeja |
| Number of patients<br>[Count(%)] |  | 376(100) | 112(29.8) | 86(22.9) | 99(26.3) | 44(11.7) | 35(9.3) |
| Age (Mean±SD) |  | 46.56±16.4 | 44.79±16.4 | 45.73±17.6 | 45.73±13.8 | 55.93±15.4 | 44.89±17 |
| Sex | Male | 207(55.1) | 69(61.6) | 47(54.7) | 50(50.5) | 20(45.5) | 21(60) |
|  | Female | 169(44.9) | 43(38.4) | 39(45.3) | 49(49.5) | 24(54.5) | 14(40) |
| Mean Duration receiving<br>HD (in months) |  | 10.1±9.5 | 8.49±6.1 | 16.22±12.9 | 7.78±5.9 | 5.68 ± 2.5 | 12.29 ± 13.4 |
| Urban/rural (pre-dialysis<br>residence) | Urban | 254(67.6) | 60(53.6) | 65(75.6) | 91(91.9) | 24(54.5) | 14(40) |
|  | Rural | 122(32.4) | 52(46.4) | 21(24.4) | 8(8.1) | 20(45.5) | 21(60) |
| Main occupation pre-dialysis | Agriculture | 65(17.3) | 27(24.1) | 5(5.8) | 14(14.1) | 12(27.3) | 7(20) |
|  | Fishing | 6(1.6) | 3(2.7) | 2(2.3) | 1(1) | 0(0) | 0(0) |
|  | Civil Servants | 100(26.6) | 22(19.6) | 31(36) | 38(38.4) | 4(9.1) | 5(14.3) |
|  | Mining | 0(0) | 0(0) | 0(0) | 0(0) | 0(0) | 0(0) |
|  | Traders | 58(15.4) | 12(10.7) | 13(15.1) | 27(27.3) | 1(2.3) | 5(14.3) |
|  | Transport | 7(1.9) | 2(1.8) | 2(2.3) | 1(1) | 2(4.5) | 0(0) |
|  | Others | 140(37.2) | 46(41.1) | 33(38.4) | 18(18.2) | 25(56.8) | 18(51.4) |

The distribution of self-reported primary renal diseases among patients in the selected centers (Table 3) showed the diverse etiologies contributing to CKD. Hypertension was identified as the most common self-reported primary renal disease, comprising 51.9% of patients across all centers. State Specialist Hospital Maiduguri reported the highest proportion of patients attributing their CKD to hypertension at 72.7%. Unknown etiology accounted for 18.6% overall, with the highest proportion at YSUTH Damaturu at 32.1%. Diabetic Kidney Disease (DKD) and Glomerulonephritis (GN) were other notable self-reported primary renal diseases, representing 8% and 5.9% of patients, respectively, across all centers. Among these, FMC Nguru reported the highest proportion of patients with DKD at 9.1% and GN at 25%. Additionally, the category “Other” encompassed 5.9% of patients, with variations among centers.

**Table 3:** Self-reported primary renal disease.

| Table 3: Self-reported primary renal disease |  |  |  |  |  |  |  |
| --- | --- | --- | --- | --- | --- | --- | --- |
| Variables |  | All centres | ysuth | umth | ssh_maiduguri | fmc_nguru | ssh_hadeja |
| Self-reported primary renal disease | GN | 43(5.9) | 4(3.6) | 14(16.3) | 9(9.1) | 11(25) | 5(14.3) |
|  | DKD | 30(8) | 6(5.4) | 8(9.3) | 6(6.1) | 4(9.1) | 6(17.1) |
|  | 'Hypertension' | 195(51.9) | 60(53.6) | 24(27.9) | 72(72.7) | 24(54.5) | 15(42.9) |
|  | Unknown | 70(18.6) | 36(32.1) | 18(20.9) | 12(12.1) | 1(2.3) | 3(8.6) |
|  | Other | 22(5.9) | 4(3.6) | 13(15.1) | 0(0) | 1(2.3) | 4(11.4) |
| Had renal biopsy | Yes | 25(6.6) | 1(0.9) | 4(4.7) | 2(2) | 16(36.4) | 2(5.7) |
|  | No | 351(93.4) | 111(99.1) | 82(95.3) | 97(98) | 28(63.6) | 33(94.3) |
| Diabetes | Yes | 35(9.3) | 6(5.4) | 11(12.8) | 5(5.1) | 5(11.4) | 8(22.9) |
|  | No | 341(90.7) | 106(94.6) | 75(87.2) | 94(94.9) | 39(88.6) | 27(77.1) |
| Hypertension | Yes | 238(63.3) | 75(67) | 42(48.8) | 65(65.7) | 32(72.7) | 24(68.6) |
|  | No | 138(36.7) | 37(33) | 44(51.2) | 34(34.3) | 12(27.3) | 11(31.4) |
| HIV | Yes | 8(2.1) | 0(0) | 5(5.8) | 3(3) | 0(0) | 0(0) |
|  | No | 368(97.9) | 112(100) | 81(94.2) | 96(97) | 44(100) | 35(100) |
| TB | Yes | 6(1.6) | 2(1.8) | 1(1.2) | 0(0) | 2(4.5) | 1(2.9) |
|  | No | 370(98.4) | 110(98.2) | 85(98.8) | 99(100) | 42(95.5) | 34(97.1) |

#### Number of CKD Patients receiving HD (CKD-HD) by Local Government Area (LGA)

The prevalence of CKD-HD varied across local government areas within the study region. In Bade Local Government Area of Yobe State, the prevalence was notably high, with 11 per 100,000 individuals on HD (7–15 per 100,000 at a 95% confidence interval) (Table 4). This was closely followed by Jakusko LGA of Yobe State with 8 per 100,000. Nguru, Damaturu, Bursari, Potiskum, Tarmuwa, and Yunusari LGAs all had a prevalence between 3 and 4 per 100,000, while all other areas had a prevalence of less than 2 per 100,000.

**Table 4:** Prevalence of HD for CKD in some selected LGAs in Yobe and Borno States.

| LGA | State | Number of Reported Cases | Estimated Population | Sample Proportion (Per 10<sup>5</sup>) | Confidence Interval(95%) |  |
| --- | --- | --- | --- | --- | --- | --- |
|  |  |  |  |  | Lower Limit | Upper Limit |
| Bade | Yobe | 35 | 321,241 | 11.0 | 7.3 | 15.0 |
| Jakusko | Yobe | 13 | 163,035 | 8.0E | 4.0 | 12.3 |
| Askira-Uba | Borno | 16 | 273,085 | 6.0E | 3.0 | 9.0 |
| Gwoza | Borno | 11 | 229,998 | 5.0E | 2.0 | 8.0 |

In Borno State, Askira/Uba had the highest prevalence of CKD-HD, with a prevalence of 6 per 100,000 individuals on dialysis, followed by Gwoza with 5 cases per 100,000 individuals. Beyond these, no other local government area in Borno State had a prevalence exceeding 3 per 100,000 individuals. For the northeastern zone of Jigawa State which constitutes eight local government areas (Auyo, Birniwa, Guri, Hadejia, Kafin Hausa, Kaugama and Kiri Kasama), the number of observed HD patients with CKD was generally low, with all areas reporting less than 2 cases per 100,000 individuals, except for Hadejia, which stood out with 5 cases per 100,000 individuals.

#### Spatial Distribution of CKD Cases

Our spatial analysis revealed marked regional disparities in CKD-HD prevalence within the surveyed states (Figure 1). The choropleth maps (Panels A, B, C, and D) indicate a higher concentration of CKD cases in Yobe and Borno states, particularly in the Bade and Askira/Uba Local Government Areas, which reported the highest incidences of 11 and 6 cases per 100,000 individuals, respectively

**Figure 1:**
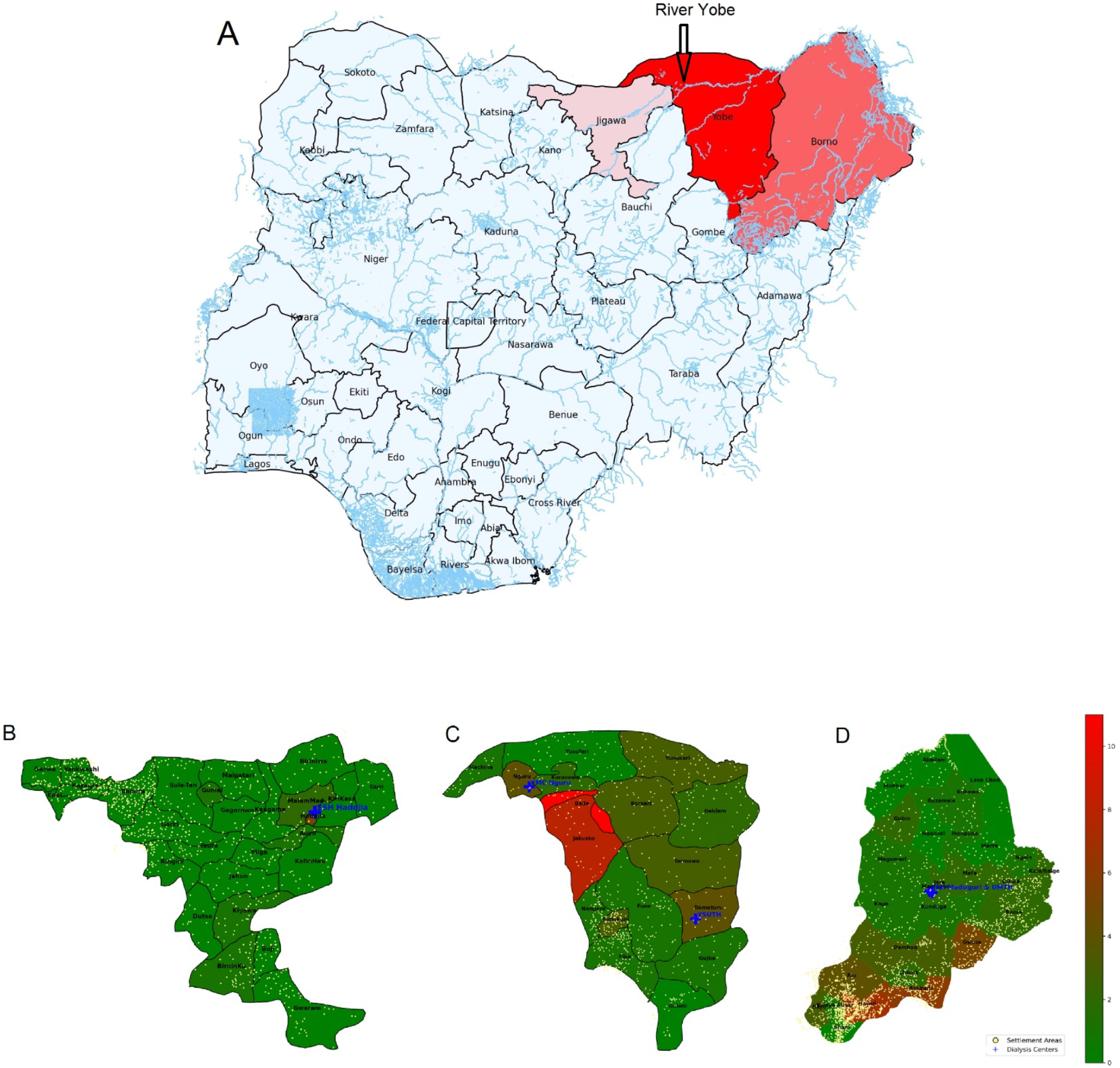
Spatial Distribution of Reported CKD-HD Cases by State and Local Government of Patient Origin. A: Choropleth Map Illustrating the Study Area and Total CKD Cases by States. B, C, & D: Choropleth Maps Depicting the Number of Reported CKD-HD Cases per hundred thousand of the individuals at the Local Level for Three States, with Yellow Points Signifying Settlement Distribution and blue crosses signifying the dialysis centers.

## DISCUSSION

We investigated the number of individuals receiving HD within the northeastern Nigeria. Focusing on some selected HD units in Yobe, Borno, and Jigawa states, our study aimed to provide a comprehensive understanding of CKD in Northeastern Nigeria. We estimated population numbers from the 2006 census data and reported population growth by WHO. ^8,9^ We collected the number of reported dialysis-dependent CKD cases in the study area and presented the distribution of CKD patients receiving HD across the local government areas of Yobe, Borno. We used HD as surrogate for CKD in the study, because a lot of CKD patients in the community may be asymptomatic and therefore may not come to hospital.

Notable findings include localized clusters of CKD patients receiving HD in the selected centers from certain local governments, such as Bade and Jakusko in Yobe State, Askira-Uba, and Gwoza in Borno State. We encountered challenges in accurately categorizing the primary renal disease (PRD) due to limitations in diagnostic facilities, especially for renal biopsy. Despite these obstacles, our findings revealed a number of cases of CKD of unknown etiology, emphasizing the urgency of enhanced healthcare infrastructure and targeted research efforts in the region.

In our study, we found that many CKD patients on HD reported hypertension as their PRD. However, it’s important to clarify that in most cases, hypertension was identified for the first time after presenting with clinical features of CKD, suggesting that hypertension was secondary to, rather than a primary cause of CKD. In addition, most patients in the study did not have their previous medical records, so we could not assess their premorbid medical history. Given the widespread presence of hypertension in the study area and its connection to kidney disease, it is possible that some of these CKD-HD cases may be unknown or secondary to a known cause such as IgA nephropathy etc since the cases were never biopsied or comprehensively investigated.

The relatively high number of HD patients in Yobe State compared to the observed rates in previous studies for West Africa, Jigawa and Kano in the northwest and even other north eastern states such as Borno and Bauchi States emphasizes the regional variability and complexity of the CKD landscape in Nigeria and may be potentially attributed to various factors such as environmental, genetic, and possible geoclimatic factors that differ across these regions. ^10–12^ It is also important to emphasize the challenges observed by Okoye and Mamven’s analysis of HD in Nigeria, where they stated that limited access to dialysis may contribute to underestimations of CKD prevalence.^13^ The relationships between our localized study and these insights emphasize the urgent need for comprehensive strategies to accurately identify and address CKD in northern Nigeria. Our study exclusively focused on patients receiving HD which may serve as surrogate for CKD; offering a limited perspective and/or a limitation on end-stage kidney disease (ESKD) within this specific cohort. It should therefore be noted that our study did not include pre-dialysis CKD, which will influence the observed prevalence and patterns of CKDu in the broader population. Furthermore, the prevalence of CKD observed among HD patients may not fully reflect the true extent of the CKD burden in the regions, as the majority of residents may face financial and geographic barriers limiting their access to HD services. Given the relatively high costs associated with HD in low-and-middle income countries (LMICs), individuals with CKD who cannot afford or access this treatment might remain undiagnosed and unaccounted for in our study. Also, factors such as cultural norms, may significantly impact women’s ability to receive dialysis.

These findings suggest the urgent need for targeted interventions, including improved access to affordable healthcare, community-based CKD screening programs, and enhanced infrastructure for early detection and management of CKD in the region.

Bade and Jakusko Local Government Areas were identified as potential hotspots, exhibiting a notably high prevalence of CKD cases. One potentially important factor may be their source of drinking water. The predominant source in these communities was borehole water, with a substantial proportion of patients using sachet water, which also originates from boreholes. This highlights the importance of investigating the quality of water, as a potential causative factor for CKD in the region. The elevated number of cases in Hadejia also raises questions about potential contributing factors. The presence of a river connecting Hadejia with northern Yobe State^14^ may facilitate the transmission of environmental contaminants or pathogens that could influence CKD prevalence. The distribution of dialysis centres and settlement patterns suggests that access to healthcare services and population density may play roles in the observed CKD distribution. This heterogeneity highlights the need for tailored healthcare interventions in the Northeastern region.

Males constitute the majority of cases in our study, with a particularly strong male presence observed at YSUTH and State SSH_Maiduguri. Conversely, SSH_Hadejia displays a more balanced gender distribution. This variance suggests the influence of distinct regional factors on CKD prevalence and aligns with previously reported region-specific trends of CKD in the north-eastern part of Nigeria.^6^ The variations may also highlight the possible lack of access to care by women. The reason why majority of CKD patients at the two HD centres in Borno State (i.e. UMTH and SSH Maiduguri) reported being resident in urban area may be due to population displacement as a result of over a decade of ‘Boko Haram’ insurgency in the region. The protracted armed conflict displaced most of the rural population in Borno state. This displaced rural population are now residing among host community and designated Internally Displaced Persons’ (IDP) camps in the capital city of Maiduguri.

## CONCLUSIONS AND RECOMMENDATIONS

In conclusion, this study sheds light on the proportions of Yobe and Borno State populations receiving HD. The spatial distribution analysis reveals potential geographic clusters of CKD cases, in specific areas, such as Bade and Jakusko Local Government areas of Yobe State. While geographic proximity and shared water resources may play a role in the observed prevalence in some regions, comprehensive epidemiological investigations are essential to understand the nature of CKD’s causative factors. In addressing the CKD challenge in northeastern Nigeria, collaboration between healthcare authorities, researchers, and communities is crucial.

To obtain a more accurate representation of CKD prevalence in the general population, comprehensive community-based surveys are crucial. Such an approach would enable the inclusion of a broader range of socioeconomic groups, providing valuable insights for public health initiatives, resource allocation, and interventions to address the wider CKD landscape in northeastern Nigeria. Environmental factors such as the source of drinking water and potential agricultural contaminants, socioeconomic disparities, limited healthcare infrastructure, climatic conditions like heat stress, and challenges in accessing timely medical services.

## Data Availability

The raw data, Images, and codes for the analysis are available https://github.com/BioRTC/CKD-HD-Survey-Paper

## Supplementary Materials

The raw data, Images, and codes for the analysis are available <u>here</u>

